# Application of a High-Resolution Melt Assay for Monitoring SARS-CoV-2 Variants in Burkina Faso and Kenya

**DOI:** 10.1101/2024.04.11.24305244

**Authors:** Caitlin Greenland-Bews, Sonal Shah, Morine Achieng, Emilie S. Badoum, Yaya Bah, Hellen C. Barsosio, Helena Brazal-Monzó, Jennifer Canizales, Anna Drabko, Alice J Fraser, Luke Hannan, Sheikh Jarju, Jean-Moise Kaboré, Mariama A. Kujabi, Maia Lesosky, Jarra Manneh, Tegwen Marlais, Julian Matthewman, Issa Nebié, Eric Onyango, Alphonse Ouedraogo, Kephas Otieno, Samuel S. Serme, Sodiomon Sirima, Ben Soulama, Brian Tangara, Alfred Tiono, William Wu, Abdul Karim Sesay, Issiaka Soulama, Simon Kariuki, Chris Drakeley, Feiko O ter Kuile, Emily R Adams, David J Allen, Thomas Edwards

**Affiliations:** The Department of Tropical Disease Biology, Liverpool School of Tropical Medicine, Liverpool, United Kingdom; Department of Infection Biology, Faculty of Infectious and Tropical Diseases, London School of Hygiene & Tropical Medicine, London, United Kingdom; Kenya Medical Research Institute, Centre for Global Health Research, Kisumu, Kenya; Groupe de Recherche Action en Santé (GRAS), Ouagadougou, Burkina Faso; Medical Research Council Unit The Gambia at London School of Hygiene and Tropical Medicine, Fajara, The Gambia; Department of Clinical Research, Faculty of Infectious and Tropical Diseases, London School of Hygiene & Tropical Medicine, London, United Kingdom; Department of Clinical Sciences, Liverpool School of Tropical Medicine, Liverpool, United Kingdom; Institut de Recherche en Sciences de la Santé (IRSS), Ouagadougou, Burkina Faso; National Heart and Lung Institute, Imperial College London, United Kingdom; Quantitative Engineering Design (QED), 30 N. Gould St., Suite 2031, Sheridan, WY 82801, USA; Faculty of Epidemiology and Population Health, London School of Hygiene & Tropical Medicine, London, United Kingdom

## Abstract

The rapid emergence and global dissemination of SARS-CoV-2 highlighted a need for robust, adaptable surveillance systems. However, financial and infrastructure requirements for whole genome sequencing (WGS) mean most surveillance data have come from higher-resource geographies, despite unprecedented investment in sequencing in low-middle income countries (LMICs) throughout the SARS-CoV-2 pandemic. Consequently, the molecular epidemiology of SARS-CoV-2 in some LMICs is limited, and there is a need for more cost-accessible technologies to help close data gaps for surveillance of SARS-CoV-2 variants. To address this, we have developed two high-resolution melt curve (HRM) assays that target key variant-defining mutations in the SARS-CoV-2 genome, which give unique signature profiles that define different SARS-CoV-2 variants of concern (VOCs). Extracted RNA from SARS-CoV-2 positive samples collected from 205 participants (112 in Burkina Faso, 93 in Kenya) on the day of enrolment in the MALCOV study (Malaria as a Risk Factor for COVID-19) between February 2021 and February 2022 were analysed using our optimised HRM assays and compared to Next Generation Sequencing (NGS) on Oxford Nanopore MinION . With NGS as a reference, two HRM assays, HRM-VOC-1 and HRM-VOC-2, demonstrated sensitivity/specificity of 100%/99.29% and 92.86/99.39%, respectively, for detecting Alpha, 90.08%/100% and 92.31%/100% for Delta and 93.75%/100% and 100%/99.38% for Omicron. The assays described here provide a lower-cost approach (<$1 per sample) to conducting molecular epidemiology, capable of high-throughput testing. We successfully scaled up the HRM-VOC-2 assay to screen a total of 506 samples from which we were able to show the replacement of Alpha with the introduction of Delta and the replacement of Delta by the Omicron variant in this community in Kisumu, Kenya. These assays are readily adaptable and can focus on local epidemiological surveillance questions or be updated quickly to accommodate the emergence of a novel variant or adapt to novel and emerging pathogens.

## Introduction

As the COVID-19 pandemic progressed, the evolution of SARS-CoV-2 gave rise to variants of concern (VOC). These VOCs posed an increased and significant threat to the global population and jeopardised public health measures and interventions that had been deployed (1). Detection and surveillance of these variants were primarily achieved through sequencing, which was crucial for tracking the spread of the VOCs worldwide. Genomic surveillance is only beneficial when representative spatially and temporally (2), and while many countries benefitted from real-time genomic surveillance during the COVID-19 pandemic, most genomic information of SARS-CoV-2 is from higher-income countries (3).

For example, as of September 2021, 18 months into the COVID-19 pandemic, sequences originating from Africa accounted for approximately 1% of the total 3.5 million sequences available (4). Similarly, it was found that as of October 2021, high-income countries were uploading 12 times more sequences than low- and middle-income countries (2). As of March 2022, there were 100,000 SARS-CoV-2 sequences available from African countries. This represented an incredible milestone in genomic surveillance in Africa and is the result of huge investments to increase sequencing capacity, with SARS-CoV-2 sequences far outnumbering any number of pathogen sequences submitted before from the continent (2). Although investments in sequencing infrastructure are ongoing, this surveillance gap highlights the need for more accessible surveillance methogs to be developed and utilised in the interim. Molecular diagnostics offer a viable alternative for targeting SARS-CoV-2 VOCs that are highly sensitive.

One promising method is high-resolution melt (HRM) assays, which feature a post-PCR analysis method that is highly sensitive in detecting nucleotide changes from shifts in amplicon melting temperature. This method has been used to identify individual mutations (5–9) with high sensitivity for detecting their respective targets. The broad range of mutations targeted across the literature includes the VOC-specific mutations N501Y, D614G, L452R, and K417N/T (7–9). However, many of these assays must be run simultaneously in singleplex to allow differentiation between multiple VOCs. This increases the work time, cost of reagents and the volume of valuable samples required for genotyping. A one-step HRM that could identify multiple mutations in one assay while cutting down on cost and time would be ideal.

Here, we build upon our previous work of one such HRM assay capable of identifying Alpha, Beta, Gamma, Delta and Omicron VOCs (10). We have developed our toolkit approach further, expanding the available primer sets targeting other VOC-defining mutations to account for previously targeted mutations now being detectable in multiple SARS-CoV-2 variants reducing confidence in distinguishing between variants. We evaluate both assays, HRM-VOC-2 and HRM-VOC-2, against NGS from Oxford Nanopore MinION, on samples collected in Burkina Faso and Kenya from February 2021 to February 2022.

## Materials and Methods

### Ethics

#### The collection of samples and their use was reviewed and approvedby the following bodies

KEMRI Scientific and Ethics Review Unit (SERU) (Ethics Reference: 4097), Kenya Pharmacy and Poisons Board, the N Health Research Ethics Committee (CERS) and Technical Committee for Clinical Trials (Comité Technique d’Examen des demandes d’autorisation d’ Essais Cliniques [CTEC]) of the Ministry of Health (Burkina Faso), the Research and Ethics Committee of Liverpool School of Tropical Medicine, (LSTM, UK) (Ethics Reference: 20-063) and the London School of Hygiene and Tropical Medicine (LSHTM, UK) (Ethics Reference: 22599).

### Sample Collection and Study Setting

All samples were collected as part of the Malaria as a Risk Factor for COVID-19 in Western Kenya and Burkina Faso (MALCOV) study (NCT04695197). Mid-nasal swabs were taken from SARS-CoV-2 positive participants and stored in viral transport media (Biocomma). Samples were collected between February 2021 and February 2022. Details of the study settings and sites involved can be found in the study protocol (11). This study was conducted across location sin the UK and sub-Saharan Africa. Assays were developed and validated in the UK, training was conducted in Kenya and Gambia, and testing was conducted in Kenya, Gambia, Burkina Faso and the UK. 112 samples from Burkina Faso and 93 from Kenya were sequenced and analysed by both HRM assays (HRM-VOC-1 and HRM-VOC-2). A further 413 samples from the Kenyan cohort were analysed by HRM-VOC-2 (Total sample count analysed by HRM-VOC-2, n=506dA) but were not sequenced, to determine the molecular epidemiology of the variants of concern.

### RNA Extraction

RNA was extracted from clinical specimens in VTM transport media using the QIAamp Viral RNA Kit (QIAGEN, Germany), following the manufacturer’s protocol and implemented as an automated workflow using the QIAcube HT platform (QIAGEN). Purified RNA was eluted in 50ul of elution buffer and stored at -80°C until use.

### RT-PCR

RT-PCR was performed by staff on site in Kenya and Burkina Faso according to the study protocol.

### Design of HRM-VOC-2 Assay

Sequences representing the known variants classified as variants of concern by the World Health Organisation (WHO), under monitoring, and of interest (VOC, VUM, and VOI, respectively) were downloaded from GenBank and aligned using ClustalX in BioEdit. Lineage-defining mutations were identified from the literature and online repository CoVariants.org (12) and located within the alignment.

Primers were designed (**Table S1**) with the aid of Primer 3 (13), and where no suitable primers could be obtained, primers were designed manually. The suitability of primers was initially tested in-silico using Oligocalc (14) and uMelt (15) to ensure compatible melting temperatures (Tms).

Singleplex testing was conducted to ensure specificity of primers and was conducted by testing each primer pair on extracted RNA from cultured viral isolates for Alpha (Genbank accession number: MW980115), Beta (hCoV-19/South Africa/KRISP-EC-K005321/2020) (BEI Resources), Gamma(hCoV-19/Japan/TY7-503/2021), Delta (SARS-CoV-2/human/GBR/Liv_273/2021), Omicron(BA.1) (SARS-CoV-2/human/GBR/Liv_1326/2021), Wild type (isolate REMRQ0001/Human/2020/Liverpool) (Alpha/Beta/Gamma/Delta/OmicronBA.1/OmicronBA.2/WT) and following this a multiplex was formed with compatible peak Tms that targeted Alpha, Delta and Omicron (BA.1).

### HRM Assays

Two multiplex HRM assays were evaluated, each containing four different primer pairs, the HRM-VOC-1 assay as described in (10) and the HRM-VOC-2 assay described above. For each assay, 2.5µl of RNA template was added for 12.5µl final reaction volumes using Lunar Universal Probe One-Step RT-qPCR kit (New England BioLabs, USA), 1X EvaGreen® dye (Biotium, USA), and primers added to their optimised concentrations (Table 1).

**Table 1:** Optimised final reaction primer concentrations for primer set in the two multiplex assays (HRM-VOC-1 and HRM-VOC-2)

| Assay | Primer set | Final Forward Primer Concentration (nM) | Final Reverse Primer Concentration (nM) |
| --- | --- | --- | --- |
| HRM-VOC-1 | S_del. 156-157 | 100 | 100 |
|  | S_K417N | 150 | 150 |
|  | N_D3L | 600 | 600 |
|  | S_EPE_HRM-VOC-1 | 250 | 250 |
| HRM-VOC-2 | S_A570D | 400 | 400 |
|  | S_L452R | 200 | 200 |
|  | S_EPE_HRM-VOC-2 | 400 | 400 |
|  | Orf_Control | 100 | 100 |

Reactions were performed using QuantStudio 5 (Thermo Fisher, US) for Kenyan Samples and QuantStudio 6/7 flex (Thermo Fisher, US) for Burkinabe samples. The thermal cycle profiles are found in Table S2.

### Analysis of HRM Assay Data

Data was visualised as negative first derivative plots using QuantStudio Design and Analysis Software (v1.5.2, QuantStudio 5 systems, (Thermo Fisher Scientific Inc).

Samples that did not yield enough sequence coverage by nanopore sequencing to identify a variant using NextClade (16) were excluded from further analysis. Samples that gave a HRM peak that could not be assigned to a variant were classified as undetermined. In the instance of HRM-VOC-2 where there is a control peak, where the control peak was absent these samples were classified as invalid. For HRM-VOC-2 if there is a control peak but the remaining peaks do not fit the signature peaks for the variants of concern and therefore can’t be assigned, these samples were classified as undetermined. For analysis of the assay performance invalid HRM results were excluded (Figures 1 and 2). Sensitivity and specificity analysis was performed in MedCalc diagnostics calculator (17).

**Figure 1:**
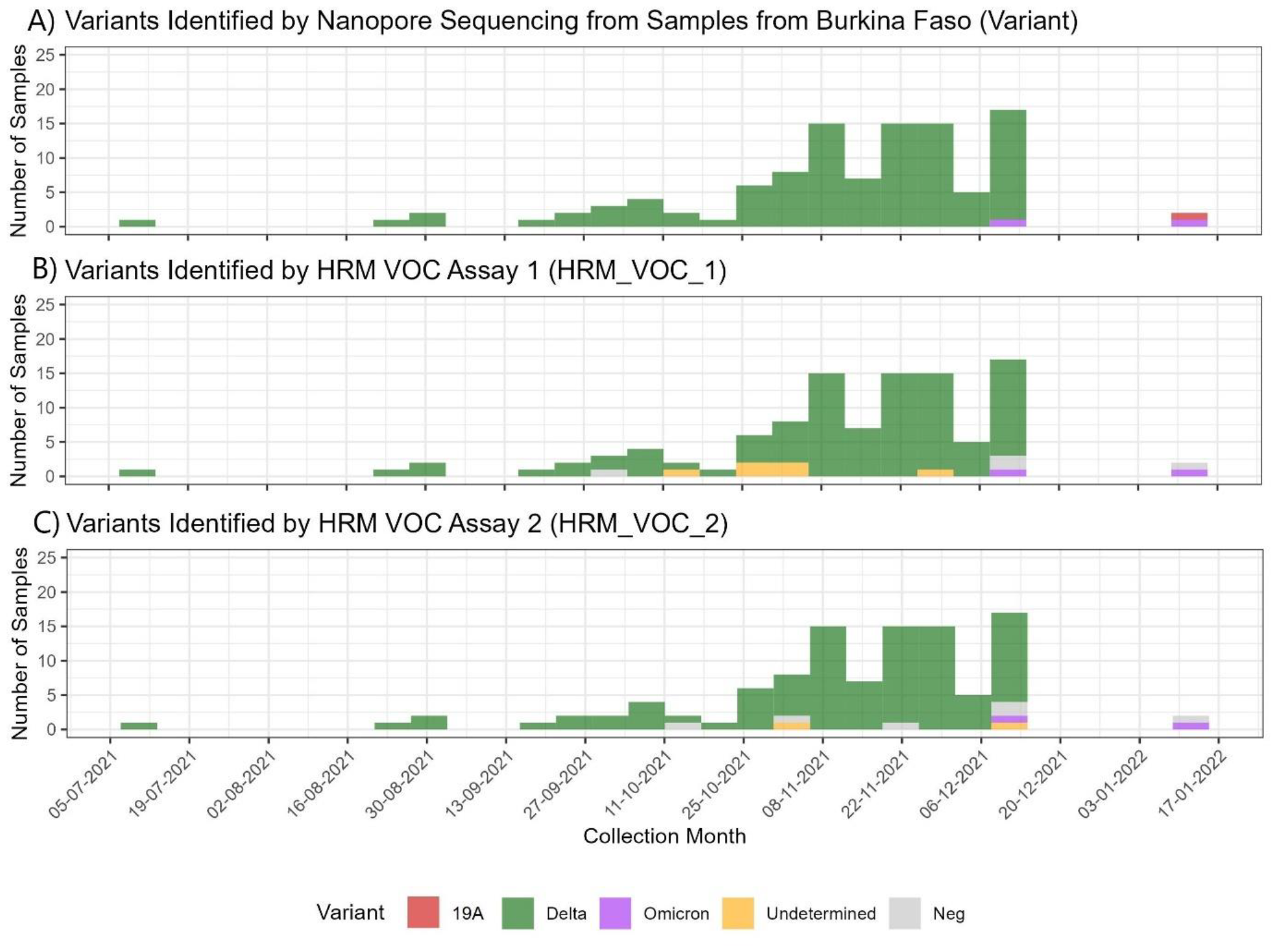
SARS-CoV-2 Variants identified in Burkina Faso using different methods. A) Number of samples collected in Burkina Faso from July 2021 to January 2022 and the variants that were identified by Nanopore Sequencing. B) Number of samples in the Burkina Faso cohort and the variant identified by using the HRM-VOC-1 assay. Negative results represent those with no amplification observed; undetermined samples had amplification but no identifiable VOC peak. C) Number of samples in the Burkina Faso cohort and the variant identified by using the HRM-VOC-2. Negative results are those where no amplification was observed, and undetermined results are those with a control peak without an identifiable VOC peak.

**Figure 2:**
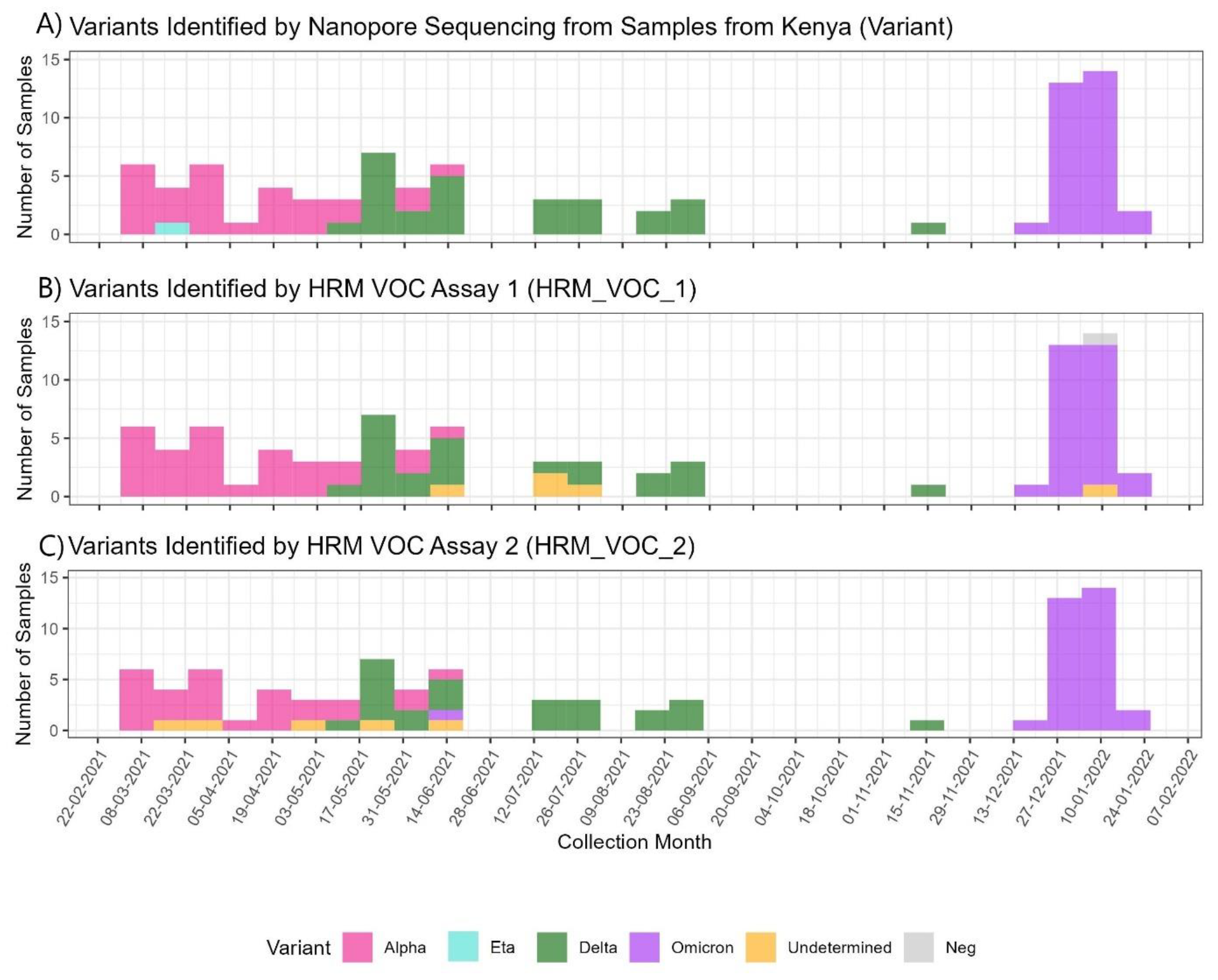
SARS-CoV-2 Variants identified in Kenya using different methods. A) Number of samples collected in Kenya from February 2021 to February 2022 and the variants that were identified by Nanopore Sequencing. B) Number of samples in the Kenyan cohort and the variant identified by using the HRM-VOC-1 assay. Negative results represent those with no amplification observed; undetermined samples had amplification but no identifiable VOC peak. C) Number of samples in the Kenyan cohort and the variant identified using the HRM-VOC-2. Negative results represent those with no amplification observed; undetermined samples had amplification but no identifiable VOC peak.

### Sequencing

205 SARS-CoV-2 samples from Burkina Faso and Kenya combined were prepared according to the Artic SARS-CoV-2 sequencing protocol (18). Amplicon generation was conducted using Artic V4.1 primers (Integrated DNA Technologies, USA). Q5® Hot Start High-Fidelity 2X Master Mix (New England Biolabs, USA), 10μM primer pools, and a thermocycling profile of 30-seconds 98 °C heat inactivation, followed by 25 cycles of 15-seconds denaturation at 98°C and 5-minute annealing/extension at 65°C. Library preparation was carried out using the Ligation Sequencing Kit (SQK-LSK109) and Native Barcoding Expansion Kits (EXP-NBD196, Oxford Nanopore Technologies, UK). Enzymes for barcode and adapter ligation were acquired from New England Biolabs (USA), and AMPure XP beads were acquired from Fisher Scientific (USA). Sequencing was performed on a R.9.4.1 flow cell on a MinION Mk1B device (Oxford Nanopore Technologies, UK) for Kenyan samples and GridION device for Burkinabè samples.

### Sequencing Analysis/Bioinformatics

Bioinformatics analysis was performed by following the Artic bioinformatics pipeline (19). Basecalling was performed using Guppy and a consensus sequence was generated. Consensus sequences were processed by NextClade (v.2.14.1) (16) for rapid variant calling and mutation summaries.

### Statistical Analysis and Data Processing

Diagnostic accuracy Samples that did not yield enough coverage from sequencing for a variant to be identified were excluded from analysis due to the lack of a reference standard.

Sensitivity, specificity and accuracy were calculated for each variant by comparison to the reference standard NGS and the calculation was performed using MedCalc (17).True positives were defined as samples where the HRM-identified variant matched the variant identified by sequencing. A true negative was every sample correctly identified as a variant other than the target VOC for that analysis. Overall agreement with the NGS result was calculated per assay as the total number of true positives divided by the total number of samples sequenced. Cohens Kappa (agreement) was calculated and interpreted per variant for each assay as described in McHugh et al (20).

### Comparison of HRM-VOC-1 and HRM-VOC-2

McNemar’s test was applied to compare the results of HRM-VOC-1 and HRM-VOC-2. This was performed for each variant (Alpha, Delta, Omicron) using the *mcnemar*.*test()* function in R.

### Data Processing and Visualisation

All data visualisation was conducted using R in RStudio (Version: 2023.3.1.446). Graphical analysis was undertaken using the ggplot2 package.

## Results

Samples were collected in Burkina Faso from July 2021 to January 2022 (Figure 1). Nanopore Sequencing identified four clades: 19A (n=1), 21I (n=67), 21J (n=37) and 21K (n=2), with most (104/112) of the samples being from one of the two Delta lineages (21I and 21J) (Figure S1). Six samples did not produce sufficient quality reads for identifying a variant using either Next Clade or Pango lineage. A total of 14 Pango lineages were identified in this dataset, with the numbers of each lineage described in the supplementary materials (Figure S1).

### Variants identified in Kisumu, Kenya

Samples were collected in Kisumu, Kenya, from February 2021 to February 2022 (F**i**gure 2). In the sample set of 93 samples nanopore sequencing identified six different clades: 20I (n=28), 21D (n=1), 21A (n=20), 21J (n=5), 21I (n=2), 21K(n=30) corresponding to four different variants, Alpha, Eta, Delta and Omicron and six Pango lineages (Figure S2). Seven samples did not yield high enough quality reads for a variant to be identified using Next Clade.

### Detection by HRM

Of the 193 samples with a valid sequencing result from both settings combined, the HRM-VOC-1 assay identified variants in 176 of these (91.2%). 118 were identified as Delta, 30 as Omicron, and 28 as Alpha. 11 samples produced a peak profile, but a variant could not be determined, and 5 samples showed no amplification and were classified as negative (Figure 2). The HRM-VOC-2 assay identified variants in 179/193 samples (92.7%), 120 were Delta variants, 33 were Omicron, and 26 were Alpha. There was one sample identified as Eta by sequencing, this was identified as false positive Alpha result by HRM-VOC-1 and gave a peak classified as ‘unidentified’ by HRM-VOC-2. Seven samples gave a positive HRM result but did not have a peak profile indicative of one of the three targeted VOCs (Alpha/Delta/Omicron), six showed no amplification and were classed as negative, and there was one invalid sample result (Figure 2). Invalid results were not included in graph visualisation or sensitivity or specificity analysis.

### Assay performance

Sensitivity and specificity were calculated for the combined HRM results across both study locations in comparison with NGS reference (Table 2). 193 samples gave a valid result when using the HRM-VOC-1 assay, and 192 samples when using the HRM-VOC-2 assay. The HRM-VOC-1 assay had a sensitivity and specificity of 100% and 99.39%, respectively, for Alpha, 90.08% and 100% for Delta and 93.75% for Omicron. The HRM-VOC-2 assay had a sensitivity and specificity of 92.86% and 99.39%, respectively, for Alpha, 92.31% and 100% for Delta and 100% and 99.38% for Omicron.

**Table 2:** Combined performance from Burkina Faso and Kenya of each HRM assay, HRM-VOC-1 and HRM-VOC-2, compared to NGS results.

| **(A) HRM-VOC-1** |  |  |  |  |  |  |  |
| --- | --- | --- | --- | --- | --- | --- | --- |
| VOC | True Positive | True Negative | False Positive | False Negative | Sensitivity(%) [CI:95 %] | Specificity(%) [CI:95 %] | Accuracy(%) |
| Alpha | 28 | 164 | 1 | 0 | 100 [87.66-100] | 99.39 [96.67-99.98] | 99.48 |
| Delta | 118 | 62 | 0 | 13 | 90.08 [83.63-94.61] | 100 [94.22-100] | 93.26 |
| Omicron | 30 | 161 | 0 | 2 | 93.75 [79.19-99.23] | 100 [97.73-100] | 98.96 |
| **(B) HRM-VOC-2** |  |  |  |  |  |  |  |
| VOC | True Positive | True Negative | False Positive | False Negative | Sensitivity(%) [CI:95 %] | Specificity(%) [CI:95 %] | Accuracy(%) |
| Alpha | 26 | 164 | 0 | 2 | 92.86 [76.50-99.12] | 99.39 [97.78-100] | 98.96 |
| Delta | 120 | 62 | 0 | 10 | 92.31 [86.31-96.25] | 100 [94.22-100] | 94.79 |
| Omicron | 32 | 159 | 1 | 0 | 100 [89.11-100] | 99.38 [96.57-99.98] | 99.48 |

**Table 3:** McNemar’s Test Result from comparing HRM-VOC-1 and HRM-VOC-2 on the combined sequenced sample set.

| Variant | McNemar's Chi Squared | df | P-value |
| --- | --- | --- | --- |
| Alpha | 1.33 | 1 | 0.25 |
| Delta | 0.08 | 1 | 0.77 |
| Omicron | 0.5 | 1 | 0.48 |

**Table 4:**
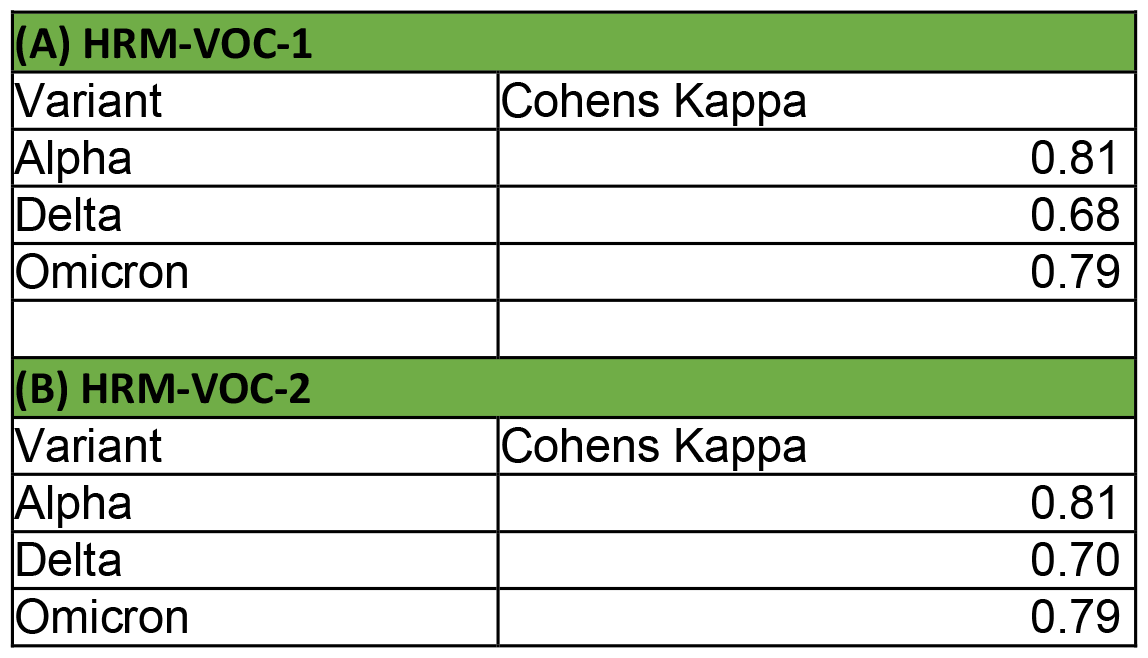
Cohens Kappa test results for comparison between HRM-VOC-1 (A) and HRM-VOC-2 (B) against sequencing results.

### McNemar’s and Cohens Kappa Test Results

No significant difference was found between the two HRM assays for detecting the three key variants using McNemar’s test (Table 3).

There was substantial agreement with the sequencing results for both HRM-VOC-1 and HRM-VOC-2, detecting Delta and Omicron, as Cohens Kappa was between 0.61-0.80, and there was almost perfect agreement with sequencing for Alpha samples (20).

### Ct Value vs Sequencing and HRM Success

All samples analysed were below RT-qPCR Ct 30, with the majority being successfully called by both assays (Figure S3). One sample gave an invalid result for HRM-VOC-1 with a Ct of 24. Nine samples gave an undetermined result for HRM-VOC-1, and 10 were undetermined by HRM-VOC-2 and had a Ct range of 23.5-29.5 in both instances.

### Scaling up Sample Screening by HRM in Kenya

Out of the 506 positive SARS-CoV-2 samples analysed by HRM-VOC-2, 396 had an identifiable variant (78.3%) (Figure 3). Of the identifiable variants, 72 samples (18.18%) were identified as Alpha, 98 samples (24.75%) were identified as Delta, and 226 samples (57.07%) were identified as Omicron. Of the remaining 110 samples, 47 gave invalid peak readings (no or limited amplification observed and absence of a control peak), 63 amplified with a control peak but the other peaks could not be categorised into signature peaks representing the variants of concern and therefore have been labelled as ‘undetermined’ (Figure 3). Cycle thresholds (Cts) were obtained from the MALCOV study team and it was determined that of these 506 samples Cts ranged from 17.9 to 39.9, with variants being successfully called across this range (Figure S5). Samples that could not be called and were labelled as invalid (Figure 3) all had a Ct of 30 or above, and undetermined samples had a range of 23.5-39.2 (Figure S5).

**Figure 3:**
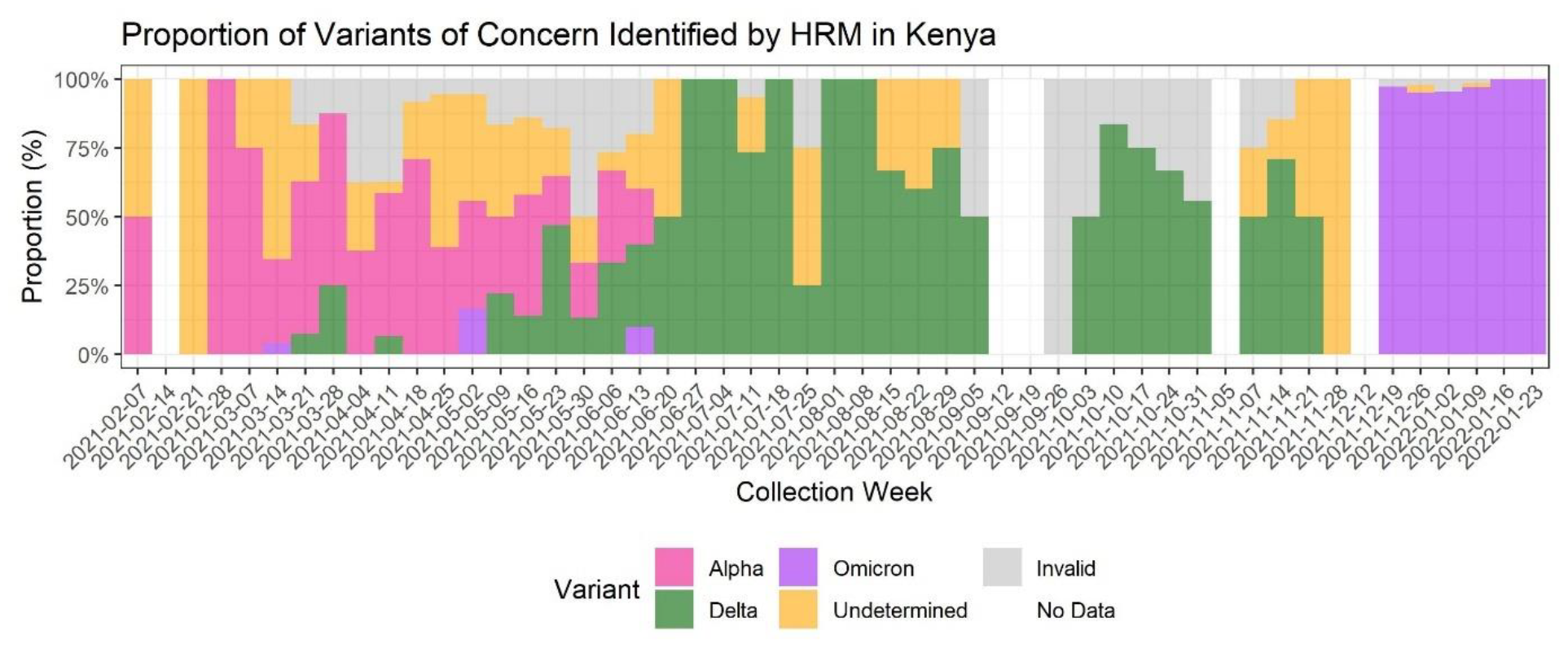
Timeseries of variants identified by HRM-VOC-2 when tested on 506 SARS-CoV-2 PCR Positive samples collected in Kenya throughout the study period. This is a combined dataset including the 86 successfully sequenced samples. Undetermined represents samples that produced a control peak but no identifiable VOC peaks. Invalid represents samples where there was no control peak. No Data represents weeks of the year where no positive samples had been collected.

Alpha was the dominant variant in the dataset at the start of sample collection (7^th^ February 2021) until early May (2^nd^ May 2021). From the 2^nd^ May 2021, the proportion of detected Delta samples increased rapidly. As of 27^th^ June 2021, Delta samples comprised 100% of samples collected. Delta remained the dominant variant detected until the 19^th^ December when Omicron fully replaced Delta at just below 100% of the total samples analysed. Three samples were identified as Omicron by HRM-VOC-2 in the first 6 months of the sample set and are represented as Omicron samples in Figure 3. Due to the timings of these samples being collected implying it is unlikely that Omicron was circulating at this time, the decision was made to label these three samples as false-positives.

## Discussion

Here we have presented the application of two variant calling HRM assays to the genotyping of positive SARS-CoV-2 samples in Burkina Faso and Kenya. These assays had high sensitivity compared to NGS for identifying all variants, with most samples being successfully variant typed by at least one assay with substantial to almost perfect agreement to NGS and no significant differences observed between the performance of the two assays.

The assay was successfully scaled up to screen more than 500 samples collected over 12 months for the MALCOV study in Kisumu, Kenya. Due to the volume of samples, it would have been expensive and labour-intensive to sequence the total sample set. However, with our HRM assay, we could identify the infecting variant in many of these samples and describe the variant waves in Kisumu during this time. Our assay has shown three variant replacement events between February 2021 and January 2022, which mirrors the three waves reported during this period from other African countries such as The Gambia (21), Ethiopia (22), and Senegal (23). In the samples analysed from Kenya this study, Alpha was the dominant variant between March 2021-May 2021 and was then replaced by Delta in May 2021, followed by Omicron in mid-December 2021, which is in keeping with epidemiological data from other regions of Kenya (24,25). Other studies have reported the Beta variant co-circulating with the Alpha variant in regions of Kenya (26). The HRM-VOC-2 assay used to screen all 506 samples in this study doesn’t detect the Beta VOC and no samples were identified as Beta in those that were sequenced, so it is unknown whether Beta was present in this sample set.

We have demonstrated that HRM is a reliable method of generating epidemiologically important data. HRM assays are also easily scalable, with 506 samples being variant-typed by the HRM-VOC-2 assay. Samples from a wide range of Ct values could be detected, however, all those invalid were above Ct 30, indicating that samples with lower Ct values should be prioritised where possible to minimise invalid results. The samples that are either unidentifiable variants or give invalid results with HRM could then be prioritised for NGS. Based on our results, this would result in only 20% of the total sample set requiring sequencing, reducing the overall expenditure. Finally, the calculated cost of this assay equates to <$1 per sample, compared to the average cost of nanopore sequencing, which has been reported to cost ∼$12 per sample when performed at high throughput (19).

Several HRM panels for variant identification have been developed throughout the pandemic. An HRM panel for detecting Delta, Omicron BA.1, BA.2 and BA.5 in four separate HRM assays achieved 97.9% agreement with Sanger sequencing (27). Another study in Iran utilised HRM for variant typing due to limited funds available for extensive sequencing and saw 93.68% sensitivity and 100% specificity compared with Sanger sequencing (6). The advantage of the approach presented here is the use of a single-tube assay that can detect Alpha, Delta and Omicron variants in one reaction without requiring multiple tests, reducing test complexity.

Throughout this study, the assays have been run on multiple instruments when used at different study sites, including QuantStudio5, QuantStudio 6/7 (Thermo Fisher Scientific Inc, USA), MIC (BioMolecular Systems, Australia) and Rotorgene Q (QIAGEN, Germany), highlighting the adaptability of the assays to multiple platforms. The transferability of this assay across platforms negates the need for instrument procurement if deciding to implement this technique, as most modern thermocyclers with the capability to perform HRM can be used.

HRM identified a small number of samples as Omicron in the first six months of the sample set. As this is a retrospective sample set, we identified these as probable false-positive results as they pre-date the established date of the first global report of Omicron BA.1 and the date of first detection in Kenya, both occurring in early November 2021 (26,28). This could be due to non-specific binding of the omicron primer sets to the RNA, or alternatively a mutation in one of the primer target sites resulting in a temperature shift of the peak that results in the shift of a peak into the Tm range for Omicron for HRM-VOC-2 resulting in the miscalling of the Omicron variant for these samples. To fully understand these false positives sequencing would need to be performed to investigate the potential mutations present in the target regions.

The main limitation of this approach, which has also been noted across the literature, is its inability to detect new, emerging mutations, as the assay design relies on pre-existing knowledge of the mutation profile of circulating variants. However, from existing WGS surveillance systems, information on novel SNPs of novel VOCs can be utilised in the design of HRMs to provide a more agile and accessible assay for more regions to have ownership of their surveillance efforts. In addition to this, unusual peaks from the HRM assays may be observed as a result of new, emerging mutations and these unusual results can act as a flag for samples to be investigated further by sequencing allowing the prioritisation of samples and avoiding overburdening of existing sequencing infrastructures.

Another limitation lies in the inter-assay variation, which can impact assay interpretation when the assay relies on small shifts in melting temperatures. To improve the analysis of HRM outputs, automation of the process could be used to reduce any user error/unreliability in peak interpretation, which could be achieved through machine learning methods that use previously analysed datasets to train an algorithm to interpret future outputs (29). This technology could be adapted to provide molecular epidemiological information on other pathogens without the expense of WGS.

The assays described here are each single-tube assays providing results in 3 hours from RNA to variant identification, making them quicker than WGS with a far more accessible and streamlined analysis. This technique can make VOC surveillance less costly and more rapid, reducing the wait time from sample to result and reducing reliance and potential over-burdening of local and external sequencing infrastructures. This assay’s high sensitivity and specificity have allowed us to investigate the molecular epidemiology of the VOC circulating in Burkina Faso and Kenya during the sample collection windows.

## Conclusion

HRM provides a quick, low-cost alternative to sequencing that can provide sensitive and specific identification of key mutations in three of the main VOCs of SARS-CoV-2: Alpha, Delta and Omicron. We have demonstrated that the assays are flexible, easily updatable and readily applied to retrospective datasets. The use of these assays would not only reduce the cost of genomic surveillance but prevent overwhelming existing sequencing infrastructure during a pandemic or outbreak situation.

## Data Availability

All sequencing data is deposited in BioProjects : PRJNA1095865 and PRJNA1096688
All other data produced in the present work can be made available upon reasonable request to the authors.

## Acknowledgements

We thank all the MALCOV study participants and the research assistants. Without them, this study would not have been feasible. We are grateful to the Directors of Health, County Health Management teams, and Medical Superintendent of Kisumu, Busia, and Siaya Counties for accommodating the study in Kenya, and the Directors of Health and staff of the Health Region of Ouagadougou, Health District of Kossodo, and the Response Centre for Health Emergencies for accommodating the study in Burkina Faso. This study is published with the permission of the Director, KEMRI. The Liverpool School of Tropical Medicine was the sponsor.

## Author Contributions

DJA, TE, CGB designed the HRM evaluation described here. DJA, TE, AJF, and CGB, designed the HRM assays. The MALCOV study was designed by FtK and CD with SS and AF as Co-PIs and HCB as country co-PI. Experimental work was conducted by DJA, TE, CGB and SS. CGB was responsible for the analysis of HRM and sequencing data for the evaluation. Experimental work in the Gambia was supervised and supported by YB, SJ, MK, JM and AKS. Experimental work in Kenya was conducted and supported by MA, BT, KO, TM and SK. Experimental work in Burkina Faso was conducted and supported by TM, SSS,IN, BIS, ESB and IS. JM. AD and WW were responsible for CRF design and development. Coordination assistance was provided by HBM and JC. EDO, AO and JMK were responsible for data management ML was senior statistician for MALCOV. LH was junior statistician for MALCOV. CGB, TE, DJA, and ERA prepared the manuscript with contributions from all authors.

## Availabilityof Data

All sequencing data is deposited in BioProjects : PRJNA1095865 and PRJNA1096688 All other data produced in the present work can be made available upon reasonable request to the authors.

## Supplementary Materials

**Figure S2:**
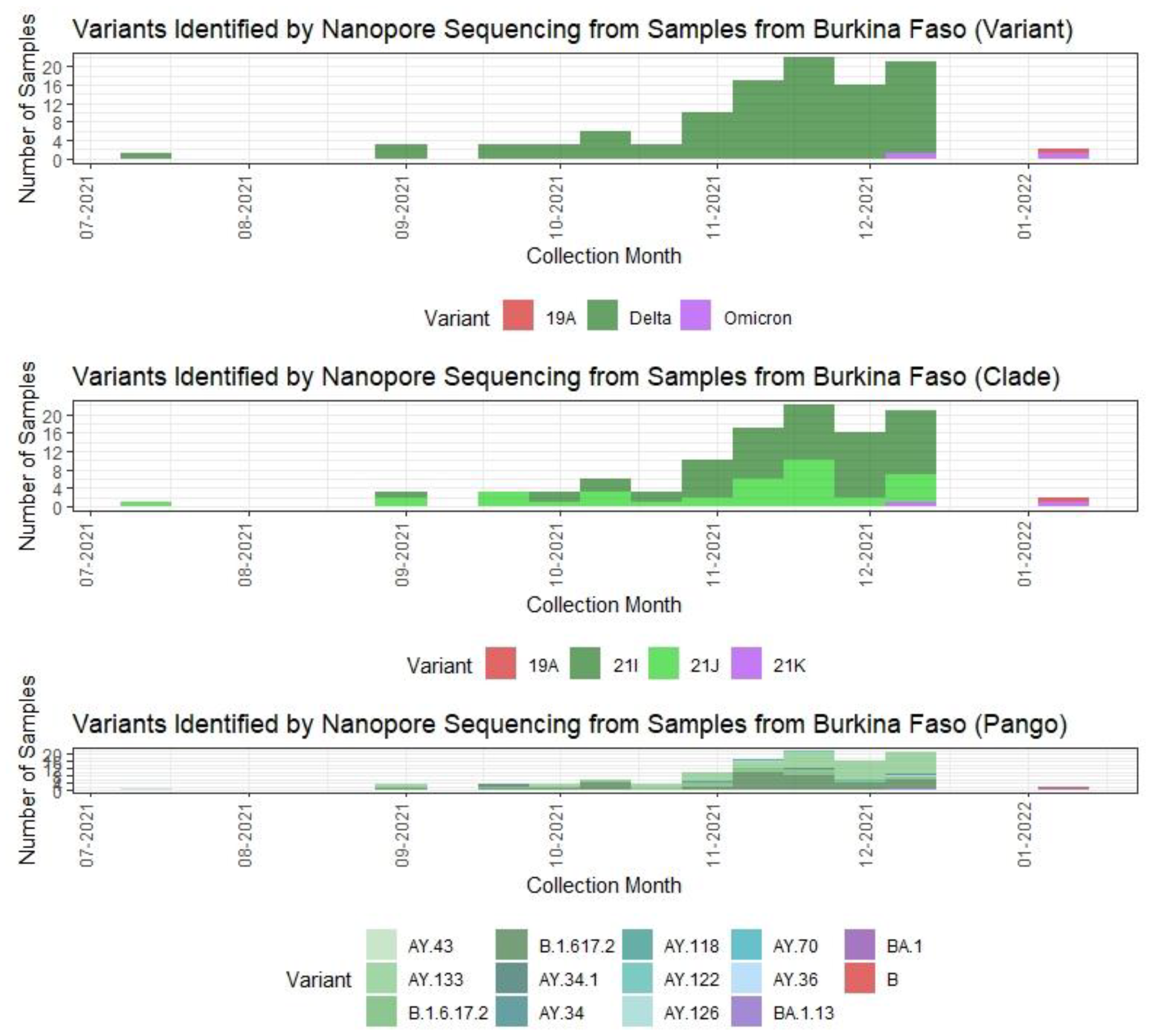
Timeseries of variants (top), Clade (middle) and Pango lineages (bottom) detected by nanopore MinION sequencing in Burkina Faso.

**Figure S2:**
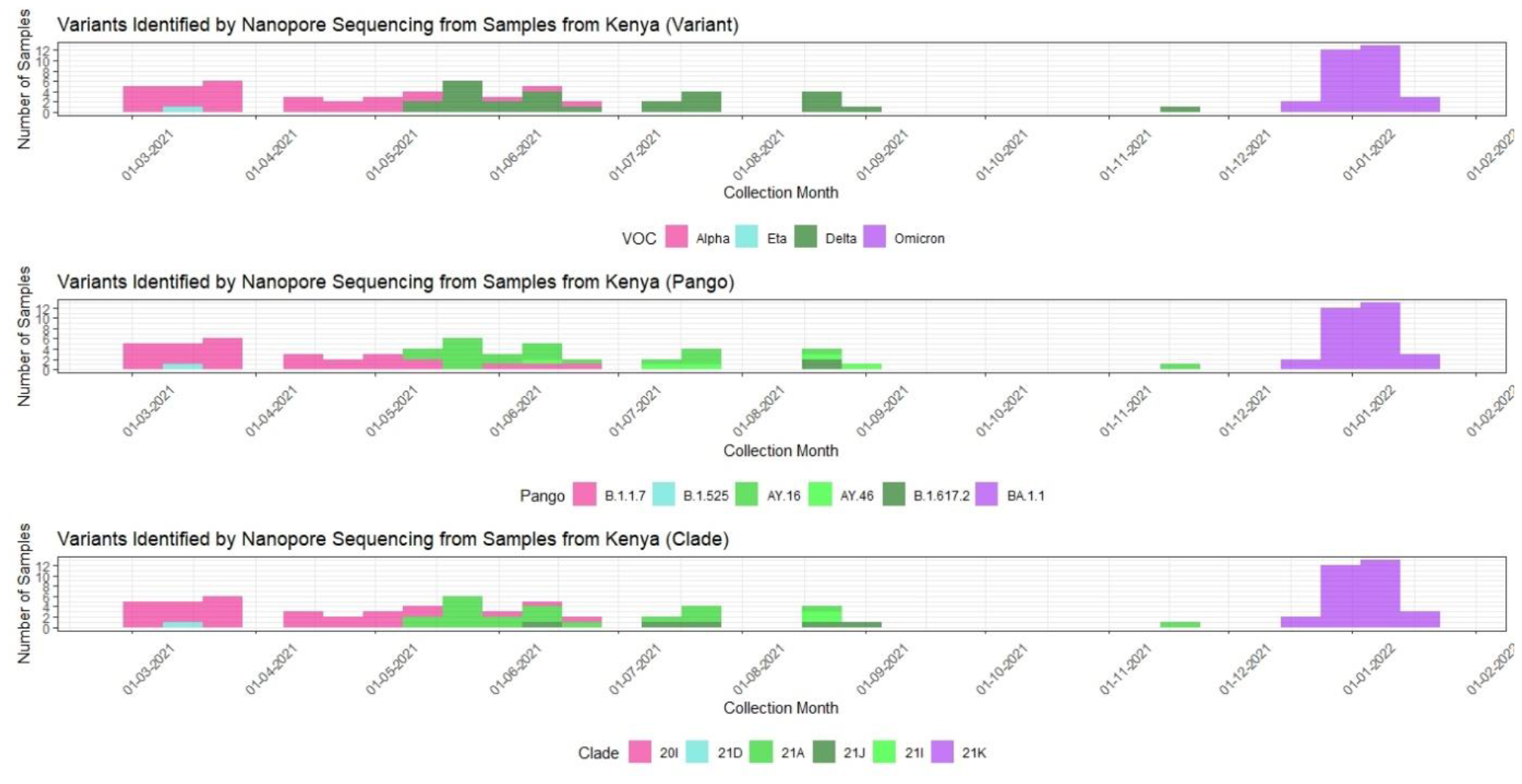
Timeseries of variants (top), Pango lineages (middle) and clades (bottom) detected by nanopore MinION sequencing in Kenya.

**Figure S3:**
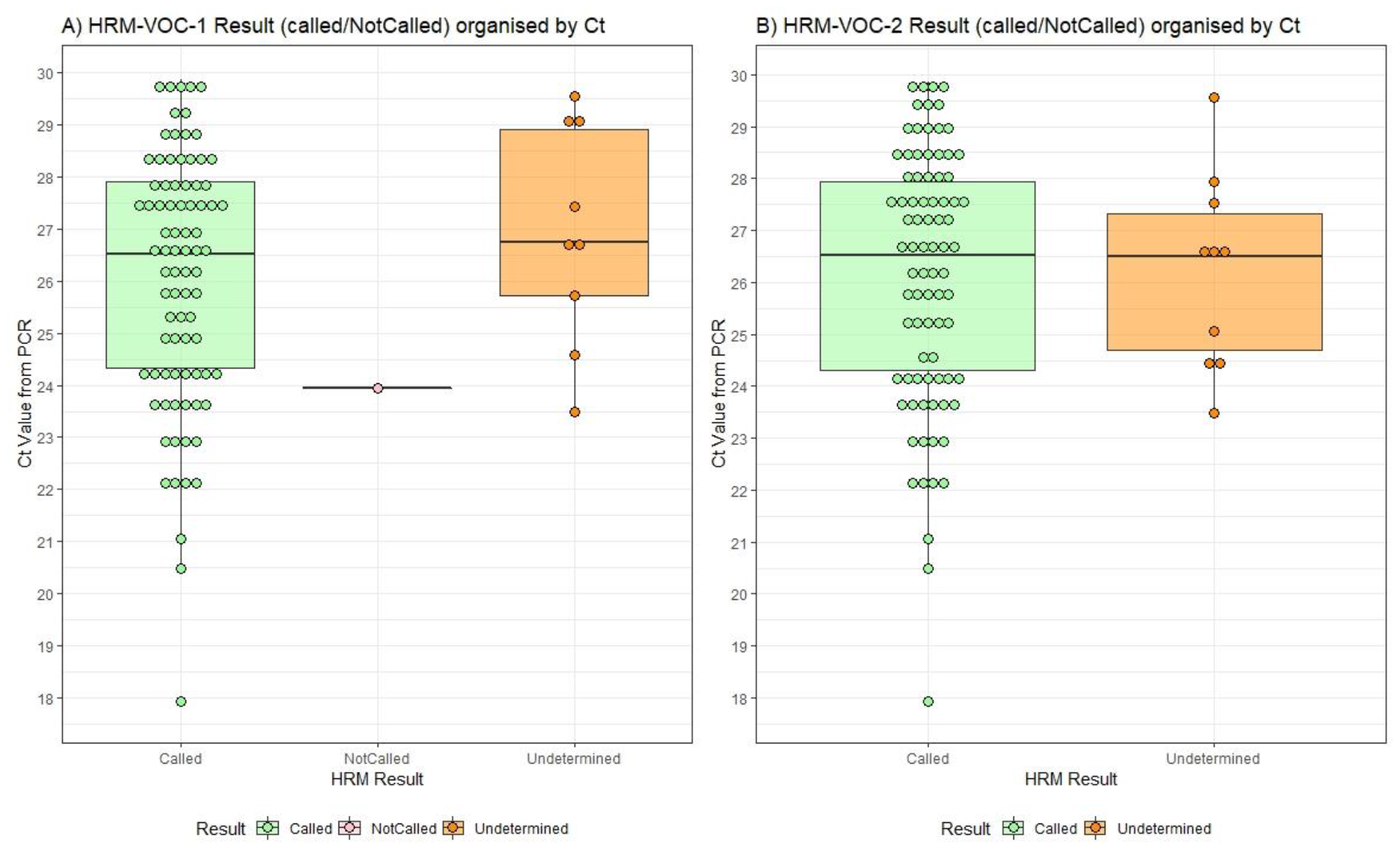
Distribution of samples by Ct from both study sites, separated into whether they could be identified as a VOC (Called), no result or invalid result (NotCalled) and where valid peaks were observed but could not be attributed to a VOC (Undetermined Variant).

**Figure S4:**
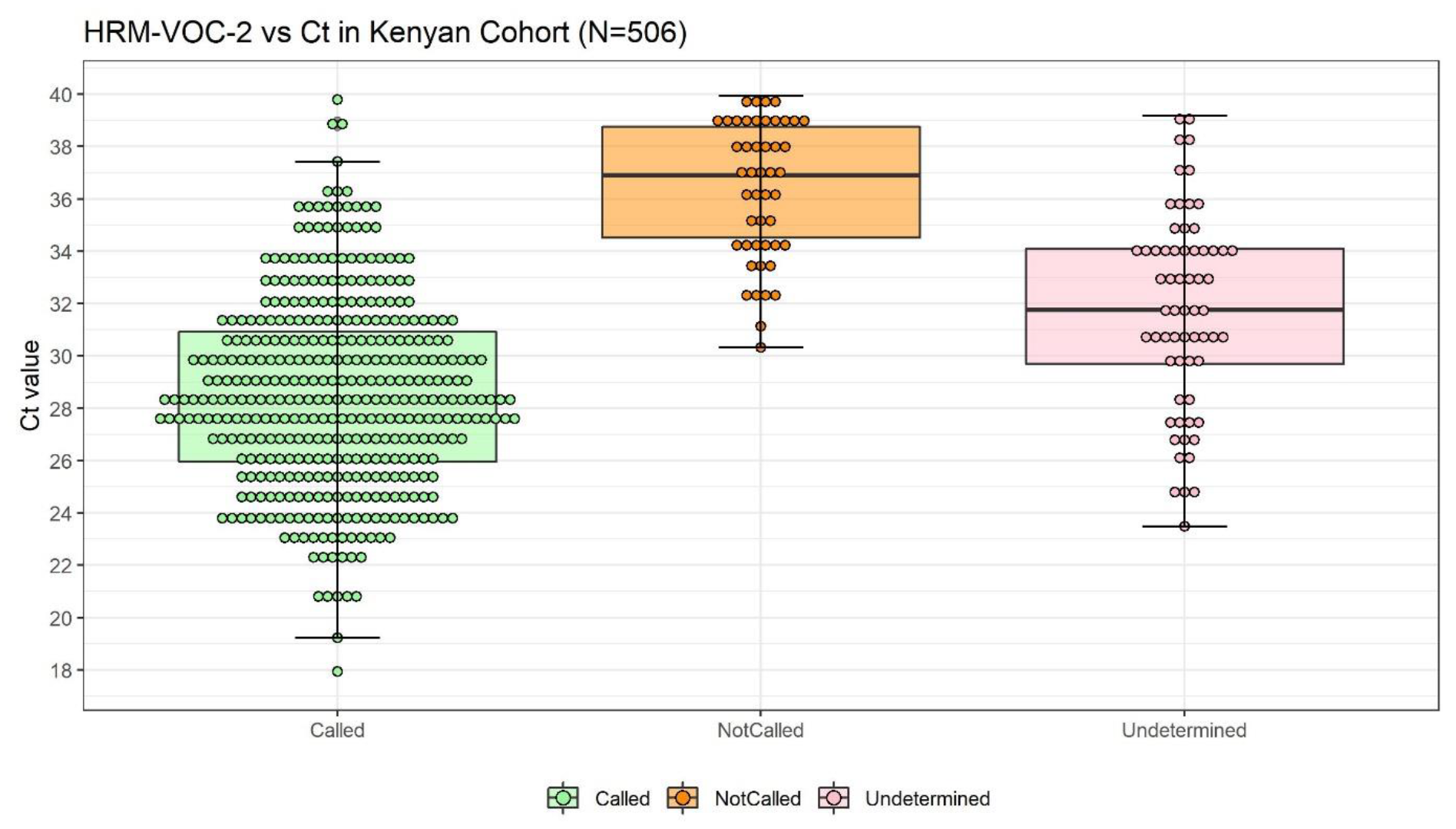
Distribution of samples by Ct from 506 samples from Kenya screened by HRM-VOC-2, separated into whether they could be identified as a VOC (Called), invalid result (NotCalled) and where valid peaks were observed but could not be attributed to a VOC (Undetermined Variant)

**Table S1:**
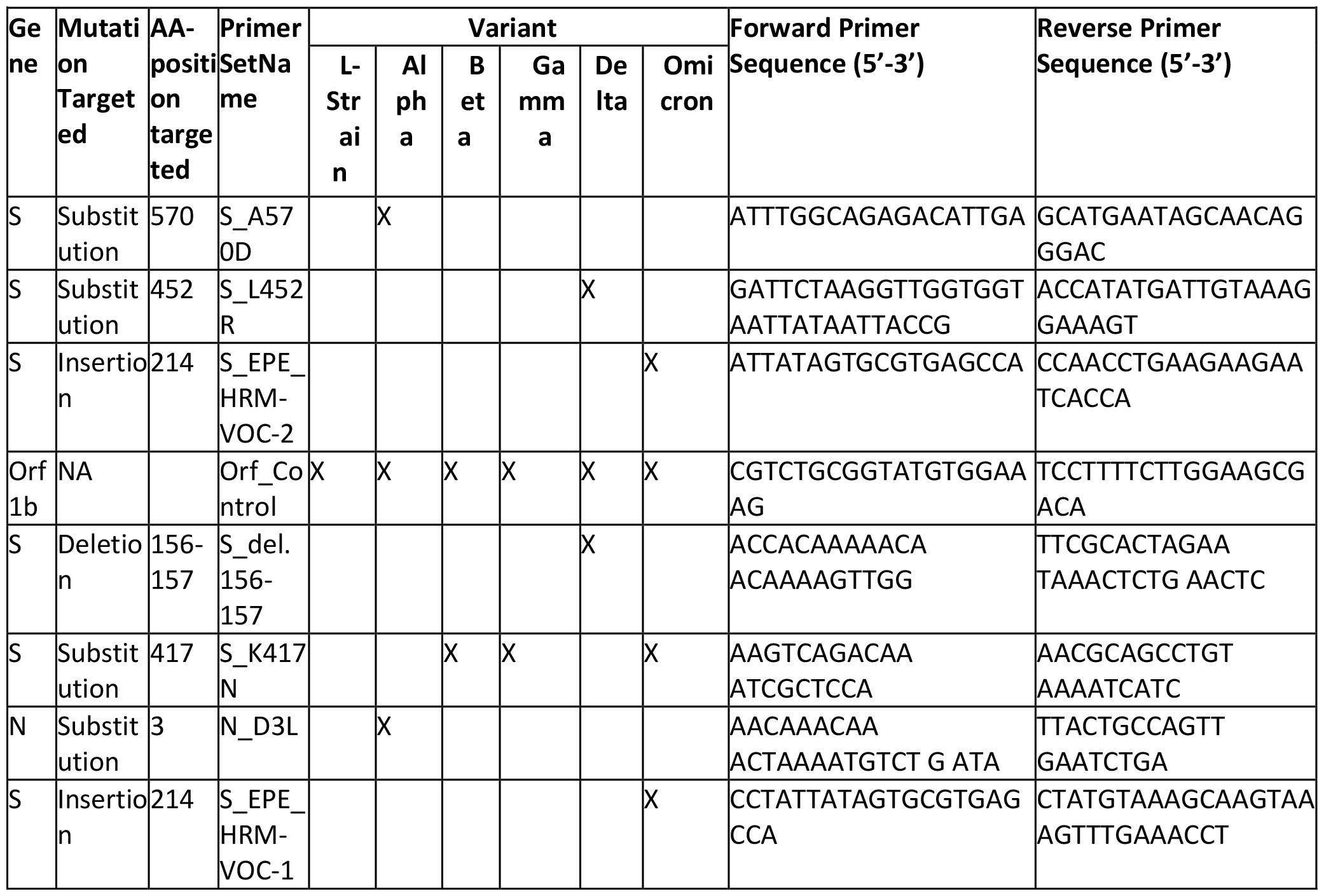
Description of the SARS-CoV-2 genome targets for, and details of the sequences of the eight primer sets designed to amplify lineage-defining mutations. These primer sets were subsequently incorporated into two multiplex assays: London and Liverpool. Orf1 = Open reading frame 1, S = Spike, N = Nucleocapsid.

**Table S2:** HRM Assay thermal cycling conditions.

| Table S2: HRM Assay thermal cycling conditions |  |  |  |  |
| --- | --- | --- | --- | --- |
| Cycling conditions: HRM-VOC-1 |  |  |  |  |
|  | Hold 1 (Reverse Transcription) | 55°C | 10min |  |
|  | Hold 2 (Initial denaturation) | 95°C | 1min |  |
| Cycling x 40 | Denaturation | 95°C | 10 secs |  |
|  | Anneal | 56°C | 30 secs |  |
|  | Extend | 72°C | 20 secs | Acquire data |
| Melt Step | HRM (0.1°C increment, 2s hold) | 73°C -85°C |  | Acquire data |
| Cycling conditions: HRM-VOC-2 |  |  |  |  |
|  | Hold 1 (Reverse Transcription) | 55°C | 10min |  |
|  | Hold 2 (Initial denaturation) | 95°C | 1min |  |
| Cycling x 38 | Denaturation | 95°C | 10 secs |  |
|  | Anneal | 60°C | 15 secs |  |
|  | Extend | 72°C | 10 secs | Acquire data |
| Melt Step | HRM (0.1°C increment, 2s hold) | 74°C -86°C |  | Acquire data |

